# Finding the Goldilocks zone for toddler accelerometry: how many days are needed for a reliable estimate of physical activity using machine learning?

**DOI:** 10.1101/2025.10.16.25338170

**Authors:** Elyse Letts, Sarah M da Silva, Sara King-Dowling, Natascja Di Cristofaro, Joyce Obeid

**Author notes:** Corresponding author: Joyce Obeid. **Data availability statement.** The datasets analyzed during the current study are available upon reasonable request to the corresponding author. **Funding statement.** This work was funded by the Canadian Institutes of Health Research. **Ethics approval statement.** This study received ethics clearance from the Hamilton integrated Research Ethics Board (HiREB #3674) and conformed to the standards of the Declaration of Helsinki. Informed written consent was obtained from a parent of each participant.

## Abstract

Accelerometers are used to measure sedentary time (SED) and physical activity (PA) in toddlers, but they may struggle to wear them for extended periods of time (e.g., weeks). Previous studies have investigated the minimum number of days needed to reliably estimate SED and PA using count-based methods. Machine learning (ML) methods use raw data which is more variable, thus potentially requiring more days for a reliable estimate. The objective of this study is to understand how many days and hours per day of accelerometer wear are needed for a reliable estimation of SED and PA using ML.

**Methods:** 109 toddlers wore an accelerometer on the right hip at home for 7 days. Time in SED, light PA (LPA), moderate-to-vigorous PA (MVPA), and total PA (TPA) were assessed using a validated ML model for toddlers. Single day intraclass coefficients (ICCs) were calculated for each minimum hours per day of wear time and each outcome. These ICCs were passed to the Spearman-Brown prophecy equation to determine the reliability of each hour per day and days combination (3-12 hours, 1-10 days).

**Results:** Predicted reliabilities ranged from 0.32 to 0.98, increasing as both numbers of hours per day and number of days increased.

**Discussion:** Our findings support the recommended 6 hours per day of wear for at least 4 days as it balances acceptable reliability with participant retention. This recommendation is valid for ML methods and we anticipate that it can be used to further explore SED and PA in toddlers using ML advances.

**Key Highlights:**

- This study calculates reliability of estimates of toddlers’ physical activity and sedentary time using machine learning method for a range of days (1-10) and hours per day (3-12).
- Our findings support the recommended 6 hours per day of wear for at least 4 days as it balances acceptable reliability with participant retention.
- We hope that this recommendation can be used to further explore physical activity and sedentary time in toddlers using machine learning advances.

## Introduction

Accelerometers are generally accepted as one of the most accurate way to measure toddlers’ (12 to 36 months of age) sedentary time (SED) and physical activity (PA) (Bruijns et al., 2020; Lettink et al., 2022), with many studies demonstrating the feasibility of wear locations including the waist (Letts, King-Dowling, et al., 2025; Trost et al., 2012), wrist (Kwon et al., 2019), and ankle (Hager et al., 2016). However, extended monitoring periods (e.g., weeks) can be difficult because toddlers may not like wearing the device(s) (Costa et al., 2013), common toddler tasks, such as naps and diaper changes, may increase device removals (Letts, da Silva, et al., 2025), and wearing devices properly relies on caregivers or early childhood educators (Costa et al., 2013; Hager et al., 2016; Kwon et al., 2019; Letts, King-Dowling, et al., 2025; Trost et al., 2012). As such, prescribed monitoring time needs to balance these practical considerations with sufficient data collection to attain reliable estimates of habitual movement behaviours.

Protocols for PA measurement with an accelerometer often ask for participants to wear the device for 7 days (Breau et al., 2022). This allows for all weekdays to be represented and so may reflect a “typical” week of activity behaviours. However, many research participants may not complete the 7 full days of wear (Breau et al., 2022). Studies have investigated the minimum number of days and hours per day required for a reliable estimate of habitual SED and PA (Addy et al., 2014; Bisson et al., 2019; Hinkley et al., 2012; Hislop et al., 2014; Kang et al., 2014; Trost et al., 2005). In adults, a minimum of 3-5 days is generally considered acceptable (Hart et al., 2011; Trost et al., 2005). In children (6-18 years) and preschoolers (3-5 years), the acceptable minimum wear ranges from 2-9 days (Addy et al., 2014; Byun et al., 2015; Hinkley et al., 2012; Hislop et al., 2014; Ramos-Munell et al., 2025). In children, the accepted minimum is 4 days (Trost et al., 2000), while in preschoolers, a minimum of 3-4 days of at least 7 hours per day (Hinkley et al., 2012; Hislop et al., 2014) is generally accepted. In toddlers (12-36 months), the current recommendations are for a minimum of 6 hours per day of wear time for at least 4 days (Bingham et al., 2016; Bisson et al., 2019; Hnatiuk et al., 2012).

All these studies, however, use count-based estimates of physical activity and sedentary time to calculate the reliability across hours and days. A count is a proprietary, unitless metric that reduces the variability of the raw accelerations data (Neishabouri et al., 2022). More recently, machine learning (ML) based methods of estimating sedentary time and physical activity have been developed that use raw accelerations as the input (Farrahi et al., 2019). A recently published ML method specific to toddlers was found to be more accurate than existing count-based methods (Letts, King-Dowling, et al., 2025). To date, no study has investigated the number of hours and days needed for a reliable measurement of SED and PA using a ML based method in toddlers. Using raw data for ML increases the variability of the data, meaning that the previous minimum wear findings, which all use counts, may not translate well to ML techniques.

Specifically, having more data variability may require more days of wear to establish a reliable estimate. As such, the aim of this study is to understand how many days and hours per day of accelerometer wear are needed for a reliable estimation of SED and PA using a toddler specific ML method.

## Methods

We recruited 109 toddlers from the Hamilton, Canada area as part the Investigating the validity and reliability of accelerometer-based measures of PhysicaL Activity and sedentarY time in toddlers (iPLAY) study. This study was approved by the Hamilton integrated Research Ethics Board (HiREB#: 3674). Toddlers were asked to wear an ActiGraph wGT3X-BT accelerometer (±8*g*; 30Hz) on the right hip on an elastic belt for 7 days, excluding sleep and water activities. Parents were also asked to record all times the accelerometer was put on and removed in a logbook.

### Accelerometer processing

Upon return of the accelerometers, ActiLife was used to download the raw data.gt3x and 1-second.agd files. The.agd files were then compared to the logbooks using a semi-automated method to determine the exact awake wear and nonwear times (Letts, da Silva, et al., 2025). Specifically, using the.agd files, we identified potential nonwear using vector magnitude counts with the following settings: a minimum nonwear length of 10 min, 0 spike tolerance, and ignored wear periods under 1 min within a larger nonwear period. Wear and nonwear periods were compared to the logbooks and corrected as needed, to ensure no nonwear periods were missed. Any naps where the accelerometer was worn were also removed from the wear time. This resulted in accelerometer data made up of only awake wear times.

Wear times were then processed in the Letts Little Movers Activity Analysis (Letts, King-Dowling, et al., 2025) tool to estimate the time spent in SED and PA. The tool reads in the raw.gt3x acceleration data and uses two distinct gradient boosted trees models to classify time spent in a) nonvolitional movement (NVM), SED, and total PA (TPA) or b) NVM, SED, light PA (LPA), and moderate-to-vigorous PA (MVPA). To do this, it first extracts 40 time and frequency domain features in 5s windows, then classifies each window as one of NVM, SED, TPA, LPA, or MVPA, and finally summarizes the daily wear time and time in each activity class. For the purposes of this study, SED was calculated by summing two activity classifications, SED and NVM (e.g., the child is sedentary as they are being pushed in a stroller or carried).

### Minimum wear time and days analysis

We estimated the reliability of measurements daily minutes of SED, LPA, MVPA, and TPA using single day intraclass correlations (ICCs) and the Spearman-Brown Prophecy equations (Trost et al., 2005). Specifically, for each outcome, we first calculated the single day ICCs for a range of minimum hours per day: 3, 4, 5, 6, 7, 8, 9, 10, 11, and 12 hours, selected based on prior studies (Bingham et al., 2016; Bisson et al., 2019; Hnatiuk et al., 2012). We filtered the data to only include days that met each of the minimum wear times being tested (3-12 hours), then fit random-intercepts linear mixed-effects models (Hislop et al., 2014) with SED/LPA/MVPA/TPA (minutes) as the outcome with participant as the grouping variable, treating days as repeated measures nested within participants. This could include any number of days per participant. The mixed-effects models provided us with the between-participant variance (σ^2^_between_) and within-participant (day-to-day) variance (σ^2^_within_). The ICC for each minimum time was then calculated (Trost et al., 2005) as:

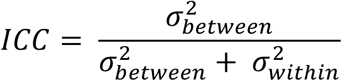

These single day ICCs for each outcome and minimum wear time were then passed to the Spearman-Brown prophecy equation (de Vet et al., 2017; Spearman, 1910; Trost et al., 2005) to calculate the projected reliability of the outcome estimations from 1 through 10 days:

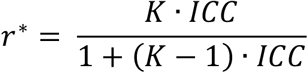

Where *r*^∗^ is the predicted reliability for *K* days.

We then summarize the estimated reliability for number of days and hours per day in heatmaps. When interpreting results, we use a reliability of 0.7 (Bingham et al., 2016; Bisson et al., 2019; Hislop et al., 2014) as the acceptable threshold.

We do not account for weekend vs weekday comparisons in our analysis. In the toddler age range, there is a much larger range of childcare arrangements, unlike in school age populations which almost exclusively have Monday to Fridays in school with weekends off. Toddlers may or may not be enrolled in daycare or other structured care settings outside of the home. If they are enrolled in daycare, the timings vary quite a lot: some children attend only half days, only on 2 days a week, etc. This variability makes it very difficult to account for. Further, previous studies in this age range have found no or minimal impact (Bingham et al., 2016; Bisson et al., 2019; Hislop et al., 2014; Penpraze et al., 2006) of weekend vs weekday vs any day inclusion. As such, we do not distinguish day of the week in this analysis.

## Results

The 109 participants (51% female) had an average age of 21.5 ± 7.0 months (range: 12.0 - 36.2 months). Participants were mostly White (72%), followed by Multiracial (20%), Asian (5%), Latin American (3%), and Arab (1%). English was the most spoken language at home (90%) and most parents were married or living with a partner (94%). Families were mostly high income (>$100,000; 52%), followed by middle income (29%), and low income (<$50,000; 17%) with 2% preferring not to disclose. Participants wore the accelerometers for an average of 6.9 ± 0.7 days (range: 3 – 9 days; any number of hours per day) and had an average daily wear time of 547.0 ± 117.3 minutes (range: 44.9 - 867.5 minutes).

Table 1 shows the ICCs for each outcome with each minimum wear time per day. ICCs range from 0.394-0.550 for SED, from 0.419-0.476 for LPA, from 0.317-0.833 for MVPA, and from 0.361-0.730 for TPA. The number of participants included in the analysis and their days of wear at each minimum daily wear time (table 1) remained relatively high until 9 hours of wear (103/109), then drops sharply at 10 hours (85/109) and beyond (11 hours: 46/109 and 12 hours: 16/109).

**Table 1.**
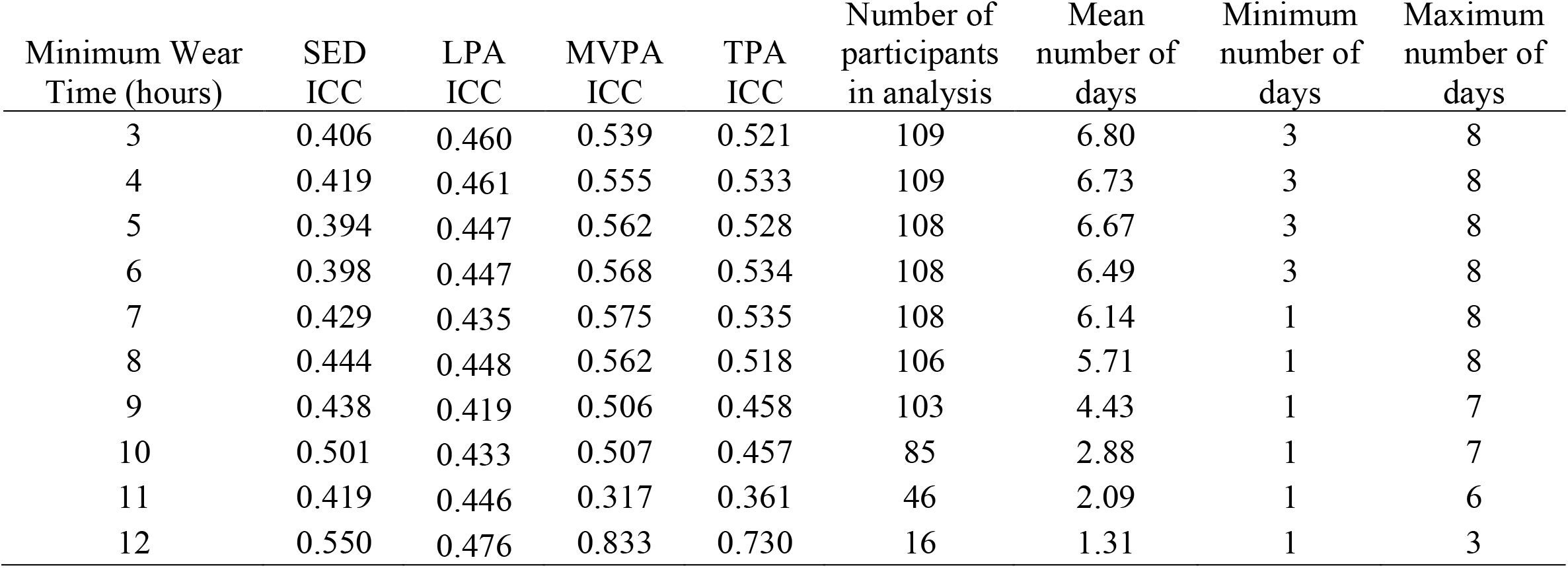
Intraclass correlations for sedentary, light physical activity, moderate-to-vigorous physical activity, and total physical activity time, including number of participants in the analysis.

Predicted reliability measures for each minimum wear time and number of days as calculated by the Spearman-Brown prophecy equation are available in Figure 1 for SED (panel A), LPA (panel B), MVPA (panel C), and TPA (panel D). Predicted reliabilities ranged from 0.32 to 0.98, generally increasing as numbers of hours per day and number of days both increased. There is, however, a decrease in reliability after the 10-hour minimum. SED and LPA tend to have lower estimated reliabilities for any given hours and days combination compared to TPA and MVPA, but still reached the 0.7 threshold.

**Figure 1.**
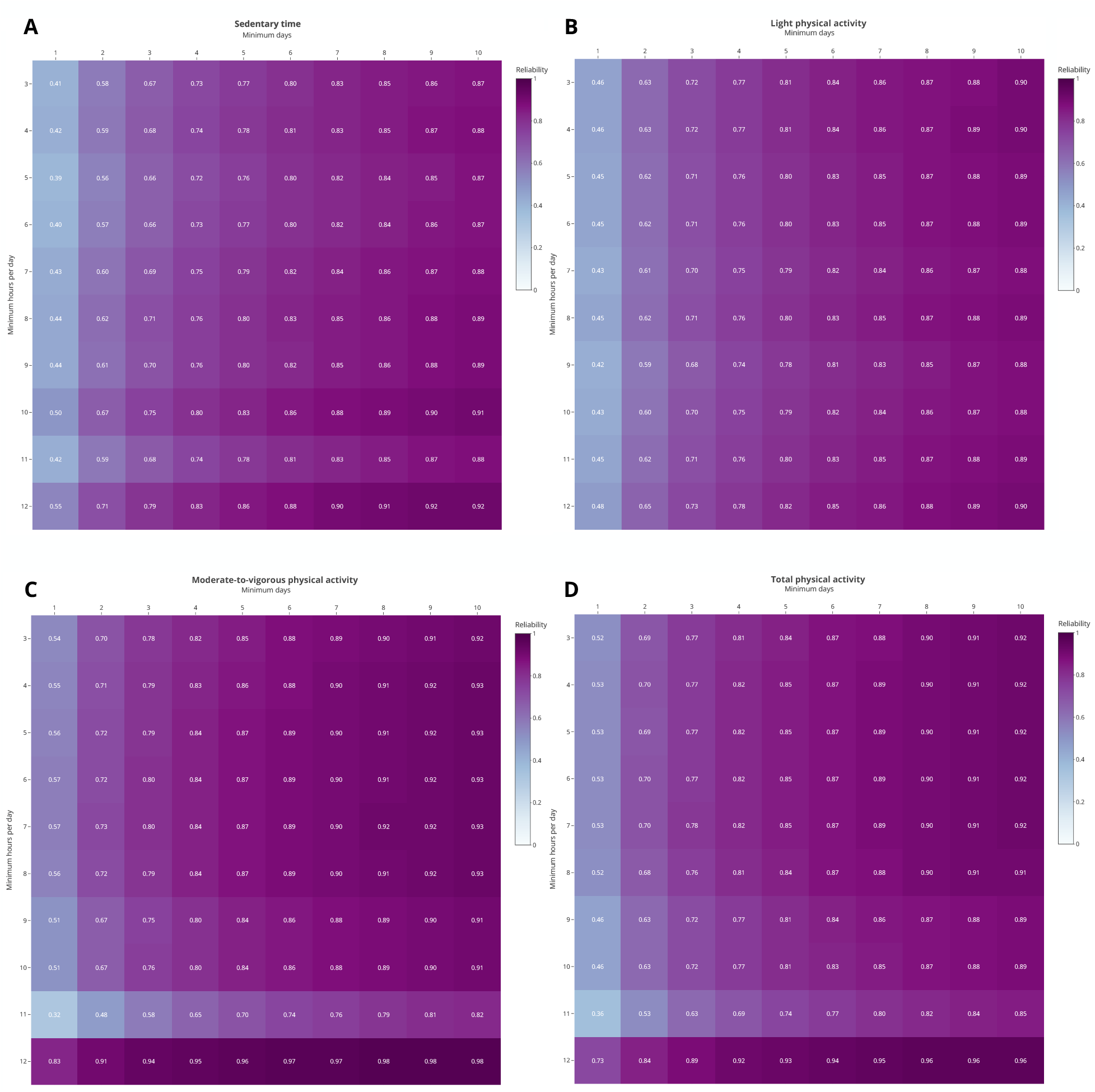
Heatmap of the predicted reliability of minimum daily wear time and number of days for sedentary time (panel A), light physical activity (panel B), moderate-to-vigorous physical activity (panel C), and total physical activity (panel D). Interactive versions of these heatmaps that allow the user to identify number of participants retained at each day and hour combination are available here: https://lettse.github.io/ToddlerMinimumDaysAccelerometry/

## Discussion

Our findings suggest that 6 hours per day of wear on at least 4 days provided acceptable reliability for estimating movement behaviours (0.72 for SED, 0.76 for LPA, 0.84 for MVPA, 0.83 for TPA). As expected, we found that the reliability of estimating movement behaviour with a toddler-specific ML method improved with increasing hours per day and number of days. In our sample, the number of participants included in the analyses remained relatively high until 9 hours per day of wear, beyond which the number of participants with valid data dropped sharply. These findings support the existing literature while providing necessary insight on a novel, ML-based approach to accelerometer data analysis.

It is not altogether surprising that reliability increased as wear time hours and days increased. Indeed, more available data results in a more representative monitoring period, which in turn translates to a more reliable measurement of free-living behaviour. However, looking at our heatmaps, a consistent pattern across all movement behaviours (SED, LPA, MVPA, TPA) emerges with a leveling-off in reliability between 9-10 hours of wear, a clear drop at 11 hours, and a sharp rebound at 12 hours. The drop between 10 and 11 hours is likely because the number of participants decreases from 78% to 42% of total participants. This aligns with previous studies that have also reported a drop after 10 hours, and likely reflects the limited amount of time toddlers spend awake on a given day (Hislop et al., 2014; Penpraze et al., 2006). The increase in reliability at 12 hours of wear is also reasonable. The 16 participants included in this analysis had fewer wear days (average of 1.31 days, maximum of 3 days), resulting in a much lower day-to-day variance (within participant variance; σ^2^_within_), which in turn leads to inflated ICCs.

Our data support previous recommendations for a minimum accelerometer wear time of 6 hours per day on at least 4 days (Bingham et al., 2016; Bisson et al., 2019; Hnatiuk et al., 2012). This combination yields reliabilities for SED, LPA, MVPA, and TPA that are above the 0.7 acceptable threshold while retaining data from 94% of participants (103/109). Selecting longer wear times would reduce the number of included participants, with 7 hours of wear excluding 10 participants (9%) and 8 hours excluding 15 participants (14%). Similarly increasing days would also reduce number of participants with 5 days (min. 6 hours) excluding 8 participants (7%). For SED, which had the lowest reliability at any given day and hour combination, the first combination to achieve a reliability of 0.8 (sometimes used as a threshold (Trost et al., 2005)) was 5 days of 8 hours of wear. However, this excludes 22 participants (20%). Further, we see these drops in participant numbers even in our sample which includes mostly White, English-speaking, higher income, and dual-parent families. Previous studies in children have shown that families with a single parent (Crumbley et al., 2024), an ethnicity other than White (Kim et al., 2023), or with a lower income (Kim et al., 2023; Wells et al., 2013) are less likely to meet minimum wear times. Therefore, more stringent minimum criteria would be likely to remove more diverse families from analyses. To balance minimum reliability while preserving the maximum number of participants and promoting diversity, we support the existing wear recommendation (Bisson et al., 2019) of 6 hours per day on at least 4 days for toddlers.

Our study is not without limitations. First, as mentioned above, we have a mostly White, English-speaking, and high-income sample which may not be representative of more diverse communities. We do however have an even split of sex and a diverse spread of ages within the toddler range. We did not investigate the effects of weekend vs weekday days. However, previous studies in this age range have found no or minimal impact (Bingham et al., 2016; Bisson et al., 2019; Hislop et al., 2014; Penpraze et al., 2006) of weekend vs weekday vs any day inclusion. Future studies could investigate the impact of daycare vs non-daycare days.

## Conclusion

This study provides a comprehensive examination of the number of days and hours per day of accelerometer wear that are needed for a reliable estimation of SED, LPA, MVPA, and TPA using a novel, toddler-specific ML method. Our findings support the recommended 6 hours per day of wear on at least 4 days as this balances acceptable reliability with participant retention. We showed that this recommendation is valid for ML methods and anticipate that it can be used to further explore movement behaviours in toddlers using ML.

## Data Availability

The datasets analyzed during the current study are available upon reasonable request to the corresponding author.

https://lettse.github.io/ToddlerMinimumDaysAccelerometry/

## Acknowledgements

We would like to thank the participants and their families for their time and effort, as well as the funders who made this research possible. We also acknowledge the efforts of Ms. Camilla Wegrzynowska who contributed to the manual cleaning of accelerometer data.

## References

Addy, C. L., Trilk, J. L., Dowda, M., Byun, W., & Pate, R. R. (2014). Assessing Preschool Children’s Physical Activity: How Many Days of Accelerometry Measurement. Pediatric Exercise Science, 26(1), 103–109. 10.1123/pes.2013-0021

Bingham, D. D., Costa, S., Clemes, S. A., Routen, A. C., Moore, H. J., & Barber, S. E. (2016). Accelerometer data requirements for reliable estimation of habitual physical activity and sedentary time of children during the early years—A worked example following a stepped approach. Journal of Sports Sciences, 34(20), 2005–2010. 10.1080/02640414.2016.1149605

Bisson, M., Tremblay, F., Pronovost, E., Julien, A.-S., & Marc, I. (2019). Accelerometry to measure physical activity in toddlers: Determination of wear time requirements for a reliable estimate of physical activity. Journal of Sports Sciences, 37(3), 298–305. 10.1080/02640414.2018.1499391

Breau, B., Coyle-Asbil, H. J., & Vallis, L. A. (2022). The Use of Accelerometers in Young Children: A Methodological Scoping Review. Journal for the Measurement of Physical Behaviour, 5(3), 185–201. 10.1123/jmpb.2021-0049

Bruijns, B. A., Truelove, S., Johnson, A. M., Gilliland, J., & Tucker, P. (2020). Infants’ and toddlers’ physical activity and sedentary time as measured by accelerometry: A systematic review and meta-analysis. International Journal of Behavioral Nutrition and Physical Activity, 17(1), 1–14. 10.1186/S12966-020-0912-4

Byun, W., Beets, M. W., & Pate, R. R. (2015). Sedentary Behavior in Preschoolers: How Many Days of Accelerometer Monitoring Is Needed? International Journal of Environmental Research and Public Health, 12(10), Article 10. 10.3390/ijerph121013148

Costa, S., Barber, S. E., Griffiths, P. L., Cameron, N., & Clemes, S. A. (2013). Qualitative Feasibility of Using Three Accelerometers With 2–3-Year-Old Children and Both Parents. Research Quarterly for Exercise and Sport, 84(3), 295–304. 10.1080/02701367.2013.812002

Crumbley, C., Cepni, A. B., Taylor, A., Thompson, D., Moran, N. E., Olvera, N., O’Connor, D. P., Johnston, C. A., & Ledoux, T. A. (2024). Exploring Factors Associated With Accelerometer Validity Among Ethnically Diverse Toddlers. Pediatric Exercise Science, 36(2), 66–74. 10.1123/pes.2022-0114

de Vet, H. C. W., Mokkink, L. B., Mosmuller, D. G., & Terwee, C. B. (2017). Spearman–Brown prophecy formula and Cronbach’s alpha: Different faces of reliability and opportunities for new applications. Journal of Clinical Epidemiology, 85, 45–49. 10.1016/j.jclinepi.2017.01.013

Farrahi, V., Niemelä, M., Kangas, M., Korpelainen, R., & Jämsä, T. (2019). Calibration and validation of accelerometer-based activity monitors: A systematic review of machine-learning approaches. Gait & Posture, 68, 285–299. 10.1016/J.GAITPOST.2018.12.003

Hager, E. R., Gormley, C. E., Latta, L. W., Treuth, M. S., Caulfield, L. E., & Black, M. M. (2016). Toddler physical activity study: Laboratory and community studies to evaluate accelerometer validity and correlates. BMC Public Health, 16(1), 936. 10.1186/s12889-016-3569-9

Hart, T. L., Swartz, A. M., Cashin, S. E., & Strath, S. J. (2011). How many days of monitoring predict physical activity and sedentary behaviour in older adults? International Journal of Behavioral Nutrition and Physical Activity, 8(1), 62. 10.1186/1479-5868-8-62

Hinkley, T., O’connell, E., Okely, A. D., Crawford, D., Hesketh, K., & Salmon, J. (2012). Assessing Volume of Accelerometry Data for Reliability in Preschool Children. Medicine & Science in Sports & Exercise, 44(12), 2436. 10.1249/MSS.0b013e3182661478

Hislop, J., Law, J., Rush, R., Grainger, A., Bulley, C., Reilly, J. J., & Mercer, T. (2014). An investigation into the minimum accelerometry wear time for reliable estimates of habitual physical activity and definition of a standard measurement day in pre-school children. Physiological Measurement, 35(11), 2213. 10.1088/0967-3334/35/11/2213

Hnatiuk, J., Ridgers, N. D., Salmon, J., Campbell, K., Mccallum, Z., & Hesketh, K. (2012). Physical Activity Levels and Patterns of 19-Month-Old Children. Medicine & Science in Sports & Exercise, 44(9), 1715. 10.1249/MSS.0b013e31825825c4

Kang, M., Bjornson, K., Barreira, T. V., Ragan, B. G., & Song, K. (2014). The minimum number of days required to establish reliable physical activity estimates in children aged 2–15 years. Physiological Measurement, 35(11), 2229. 10.1088/0967-3334/35/11/2229

Kim, E. H., Jenness, J. L., Miller, A. B., Halabi, R., de Zambotti, M., Bagot, K. S., Baker, F. C., & Pratap, A. (2023). Association of Demographic and Socioeconomic Indicators With the Use of Wearable Devices Among Children. JAMA Network Open, 6(3), e235681. 10.1001/jamanetworkopen.2023.5681

Kwon, S., Zavos, P., Nickele, K., Sugianto, A., & Albert, M. V. (2019). Hip and wrist-worn accelerometer data analysis for toddler activities. International Journal of Environmental Research and Public Health, 16(14), 2598. 10.3390/ijerph16142598

Lettink, A., Altenburg, T. M., Arts, J., van Hees, V. T., & Chinapaw, M. J. M. (2022). Systematic review of accelerometer-based methods for 24-h physical behavior assessment in young children (0-5 years old). The International Journal of Behavioral Nutrition and Physical Activity, 19(1), 116. 10.1186/s12966-022-01296-y

Letts, E., da Silva, S. M., Di Cristofaro, N., King-Dowling, S., & Obeid, J. (2025). Beyond the (Log)book: Comparing Accelerometer Nonwear Detection Techniques in Toddlers. Child: Care, Health and Development, 51(4), e70133. 10.1111/cch.70133

Letts, E., King-Dowling, S., Cristofaro, N. D., Tucker, P., Cairney, J., Kobsar, D., Timmons, B. W., & Obeid, J. (2025). Development and accuracy of a novel machine learning model to detect toddlers’ physical activity and sedentary time using accelerometers: Little Movers Activity Analysis. medRxiv. 10.1101/2025.04.24.25326266

Neishabouri, A., Actigraph, J. N., Samuelsson, J., Actigraph, T. G., Biggs, M., Jeremy, A., Actigraph, W., Cross, D., Karas, A. M., Hidalgo, J., Karolinska, M., Khan, I. S., Cong, C., & Actigraph, G. (2022). Quantification of Acceleration as Activity Counts in ActiGraph Wearables. Scientific Reports, 12. 10.1038/s41598-022-16003-x

Penpraze, V., Reilly, J. J., MacLean, C. M., Montgomery, C., Kelly, L. A., Paton, J. Y., Aitchison, T., & Grant, S. (2006). Monitoring of Physical Activity in Young Children: How Much Is Enough? Pediatric Exercise Science, 18(4), 483–491. 10.1123/pes.18.4.483

Ramos-Munell, J., Antczak, D., Álvarez-Barbosa, F., Alfonso-Rosa, R. M., Cruz, B. del P., & Pozo-Cruz, J. del. (2025). Accelerometer Monitoring Duration for Reliable Estimates of Physical Activity, Sedentary Behavior, and Step Counts in Preschoolers. Journal of Physical Activity and Health, 1(aop), 1–8. 10.1123/jpah.2024-0640

Spearman, C. (1910). Correlation calculated from faulty data. British Journal of Psychology, 3(3), 271–295.

Trost, S. G., Fees, B. S., Haar, S. J., Murray, A. D., & Crowe, L. K. (2012). Identification and validity of accelerometer cut-points for toddlers. Obesity, 20(11), 2317–2319. 10.1038/oby.2011.364

Trost, S. G., Mciver, K. L., & Pate, R. R. (2005). Conducting Accelerometer-Based Activity Assessments in Field-Based Research. Medicine & Science in Sports & Exercise, 37(11), S531. 10.1249/01.mss.0000185657.86065.98

Trost, S. G., Pate, R. R., Freedson, P. S., Sallis, J. F., & Taylor, W. C. (2000). Using objective physical activity measures with youth: How many days of monitoring are needed? Medicine and Science in Sports and Exercise, 32(2), 426–431.

Wells, S. L., Kipping, R. R., Jago, R., Brown, J., Hucker, D., Blackett, A., & Lawlor, D. A. (2013). Characteristics associated with requested and required accelerometer wear in children. BMJ Open, 3(8), e003402. 10.1136/bmjopen-2013-003402

